# Automated Derivation of Diagnostic Criteria for Lung Cancer using Natural Language Processing on Electronic Health Records: A pilot study

**DOI:** 10.1101/2024.02.20.24303084

**Authors:** Andrew Houston, Sophie Williams, William Ricketts, Charles Gutteridge, Chris Tackaberry, John Conibear

## Abstract

**Background:** The digitisation of healthcare records has generated vast amounts of unstructured data, presenting opportunities for improvements in disease diagnosis when clinical coding falls short, such as in the recording of patient symptoms. This study presents an approach using natural language processing to extract clinical concepts from free-text which are used to automatically form diagnostic criteria for lung cancer from unstructured secondary-care data.

**Methods:** Patients aged 40 and above who underwent a chest x-ray (CXR) between 2016-2022 were included. ICD-10 and unstructured data were pulled from their electronic health records (EHRs) over the preceding 12 months to the CXR. The unstructured data were processed using named entity recognition to extract symptoms, which were mapped to SNOMED-CT codes. Subsumption of features up the SNOMED-CT hierarchy was used to mitigate against sparse features and a frequency-based criteria, combined with univariate logarithmic probabilities, was applied to select candidate features to take forward to the model development phase. A genetic algorithm was employed to identify the most discriminating features to form the diagnostic criteria.

**Results:** 75002 patients were included, with 1012 lung cancer diagnoses made within 12 months of the CXR. The best-performing model achieved an AUROC of 0.72. Results showed that an existing ‘disorder of the lung’, such as pneumonia, and a ‘cough’ increased the probability of a lung cancer diagnosis. ‘Anomalies of great vessel’, ‘disorder of the retroperitoneal compartment’ and ‘context-dependent findings’, such as pain, statistically reduced the risk of lung cancer, making other diagnoses more likely. The performance of the developed model was compared to the existing cancer risk scores, demonstrating superior performance.

**Conclusions:** The proposed methods demonstrated success in leveraging unstructured secondary-care data to derive diagnostic criteria for lung cancer, outperforming existing risk tools. These advancements show potential for enhancing patient care and results. However, it is essential to tackle specific limitations by integrating primary care data to ensure a more thorough and unbiased development of diagnostic criteria. Moreover, the study highlights the importance of contextualising SNOMED-CT concepts into meaningful terminology that resonates with clinicians, facilitating a clearer and more tangible understanding of the criteria applied.

## Background

Lung cancer stands as one of the most common and serious types of cancer, ranking 2^nd^ in terms of new cases and 1^st^ in terms of mortalities, according to global statistics from 2020 [1]. The most recent statistics show that, in England, only 29.4% of lung cancer cases are identified at stages 1 and 2 [2], underscoring the critical need for improved diagnostic criteria and detection methods to enhance the chances of successful treatment and reduce the burden of this disease on patients and healthcare systems. Recognising this urgency, the NHS has a long-term plan to diagnose 75% of all lung cancers at stages 1 and 2 by 2028, aiming to significantly improve early detection rates and patient outcomes.

Early diagnosis is imperative given the aggressive nature of lung cancer, with delays in detection resulting in patients presenting with more advanced stages of the disease. Recent data published by the Office for National Statistics and Public Health England showed the 5-year survival rate among patients diagnosed with stage 1 lung cancer was 56.6%, with this figure reducing to only 2.9% among those diagnosed with stage 4 disease [3]. Additionally, precise diagnostic criteria play a pivotal role in distinguishing lung cancer from a spectrum of cardiothoracic and respiratory conditions that may exhibit similar symptoms. With that said, to ensure the cost-effectiveness and cost-benefit of targeted interventions aimed at improving the diagnosis of lung cancer, judicious allocation of resources is required [4].

Electronic health records (EHRs) have revolutionized clinical research by offering a vast and comprehensive repository of patient information. Records encompass a range of data, including patient demographics, medical history, laboratory results, medication prescriptions, and procedure information. Such extensive and structured data enable researchers to conduct large-scale, population-based studies, aiding in the identification of trends [5], risk factors [6–8], and treatment outcomes [9,10]. However, a significant limitation of EHRs pertains to accuracy and completeness, particularly among symptoms and diagnosis data. Symptoms and diagnoses are often documented in unstructured free-text clinical notes, requiring manual coding into clinical ontologies such as ICD-10 and SNOMED-CT. The process of clinical coding can introduce inaccuracies and ’missingness’ in the data, posing considerable challenges for clinical research [11,12]. Considering these challenges, techniques such as natural language processing (NLP) offer valuable solutions for not only mitigating the limitations of structured data but also unlocking valuable insights that may be exclusive to free-text narratives of patient encounters.

Natural Language Processing (NLP) has gained utility in extracting and analysing information in Electronic Health Records (EHRs). Koleck et al. (2019) conducted a literature review, finding 27 relevant studies using NLP to analyse symptoms in EHR narratives [13]. NLP has been used for auditing discharge reports [14], predicting readmissions [15], and aiding in diagnosis [16–18]. Weissman et al. (2016) used NLP to classify discharge documents based on critical illness-related keywords with high accuracy [14]. Greenwald et al. (2017) developed an NLP tool to extract readmission-related concepts and achieved comparable performance to existing prediction models [15]. In oncology, NLP extracted features from CT reports for predicting lymph node metastasis in non-small cell lung cancer (NSCLC) with competitive performance [16]. Despite its potential, there’s a gap in applying NLP to oncology symptoms, highlighting an opportunity for further research [13].

While NLP has demonstrated its effectiveness in various healthcare applications, there is a growing recognition of the advantages of extracting ontological concepts rather than use-case-specific concepts [19,20]. This approach provides a more generalised framework for understanding and organising medical information, contributing to interoperability [21,22] and facilitating the linkage with already coded, structured, clinical data found in the EHR. This transition to ontological concept extraction aligns with the broader adoption of standardised medical terminologies like SNOMED CT, which play an important role in structuring and organizing clinical data for improved healthcare decision-making and research.

From a machine learning perspective, extraction of concepts from a hierarchical ontology offers a crucial advantage, enabling the retention of valuable information, even when a patient reports rarer or more specific symptoms. For instance, when a patient mentions a symptom like a ‘chesty cough’, machine learning systems can link it to a higher-level concept in the ontology, such as “cough.” This hierarchical relationship allows the model to preserve the broader context and meaning of the symptom, preventing the loss of nuanced information that might occur in non-hierarchical concept lists, where rare features might otherwise be removed. Failure to account for such sparsity could result in poor or unreliable classification performance [23,24].

Given the promise of NLP for the accurate extraction of relevant features, at scale, this study applies NLP to extract SNOMED-CT concepts from free-text notes, applies subsumption to elevate rarer symptoms up the ontological hierarchy, then feeds the final feature set into a machine learning framework to train a model to discriminate lung cancer from other diseases. Furthermore, this study provides an exploration into how feature weights might be affected by demographic information like age, sex and ethnicity.

## Methodology

### Eligibility

Data were extracted from the Barts Health NHS Trust Data Warehouse for all patients meeting the following eligibility criteria: Patients referred for a chest x-ray (CXR), aged 40 years or older at the point of referral, during two time periods between 01 Jan 2016 and 31 Dec 2019 or 01 Jan 2022 and 31 Dec 2022 were eligible for inclusion. The time window of 01 Jan 2020 - 31 Dec 2021 were not considered due to deviations from the typical cancer care-pathways as a result of the COVID-19 pandemic. Patients who had opted out of their data being used for research, those without medical notes beyond four years from the original x-ray, unless a second confirmatory x-ray within four years ruled out lung cancer, and patients with an existing or historical diagnosis of any cancer were excluded from participation in the study.

### Data Sources

All free-text data contained in the secondary care EHR system, from one year prior to the date of the first chest X-ray, were extracted and combined with demographic information, including Age, Sex and Ethnicity, and ICD-10 data from the same time period. Additionally, diagnostic data in the form of ICD-10 codes and the Somerset Cancer Registry were extracted for the subsequent four years post-CXR, or up to the maximum available timepoint.

To determine the ground truth, a patient was labelled as having lung cancer if a diagnosis was recorded in the Somerset Cancer Registry, or an ICD-10 code of C34 (Malignant neoplasm of bronchus and lung) was present in the patient’s EHR post-CXR. Considering the potential delays in diagnoses, post-CXR, and the delays in uploading this information onto the electronic health records system, model training was performed iteratively, each time re-labelling the ground truth to consider an additional month of diagnoses. The iterative process was performed first considering only patients diagnosed with lung cancer within the subsequent month following their CXR, continuing to add more patients until 12-months post-CXR. Instances of lung cancer diagnoses over time the respective model performance is presented in Fig. 1a.

**Figure 1:**
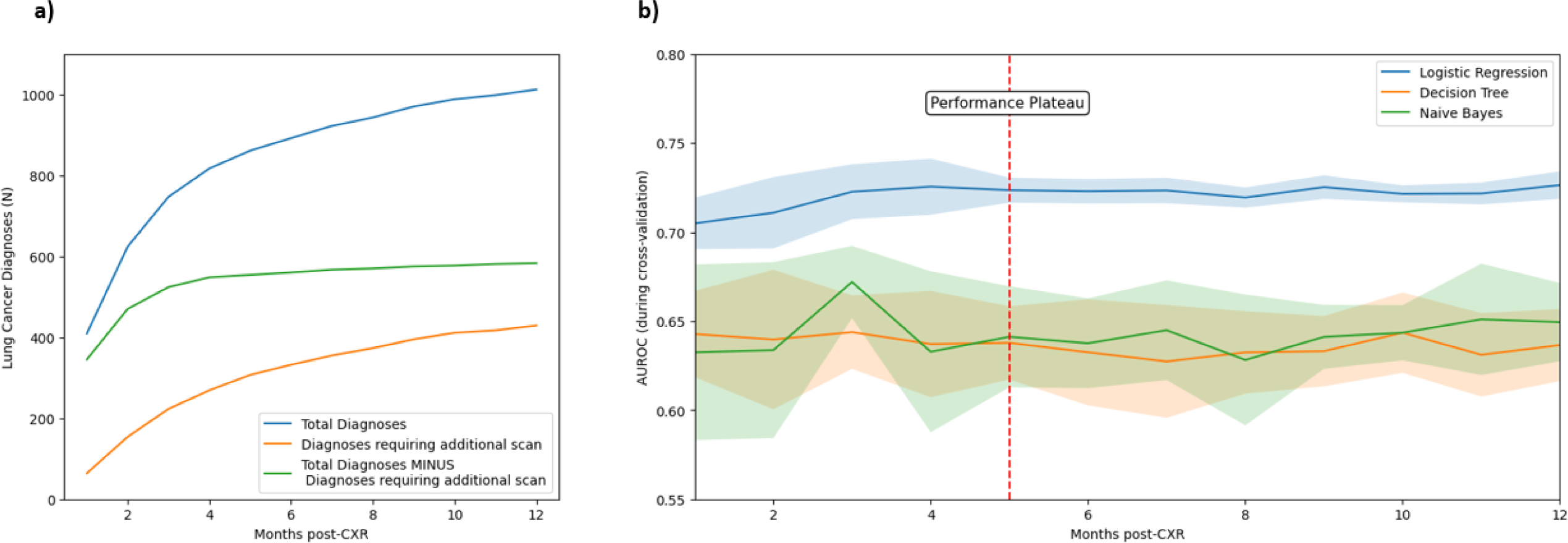
a) The number of diagnoses occurring at each monthly interval, for the subsequent 12 months post-CXR. b) The mean AUROC of the three tested models across each monthly interval, demonstrating the performance stabilisation and plateau from month five onwards. The shaded area indicates the standard deviation of the AUROC across the cross-validation folds.

### Feature Extraction

To extract structured information from the free-text data, named entity recognition (NER) was performed using the NLP software, CLiX (Clinithink Ltd., London, UK). The free-text was queried against two resource sets, a ‘Core-Problems’ list containing common clinical symptoms and diagnoses, and the Human Phenotype Ontology. The top 100 clinical features for each resource set are presented in the supplementary file.

### Feature Engineering

To handle missing data, sex and ethnicity were imputed using the most common category. Symptom data were binary, and an assumption was made that if a diagnosis or symptom was not found in either the structured ICD-10 data or identified by the NLP algorithm, the patient did not have the diagnosis or symptom.

To ensure a harmonised dataset, all features were mapped to the SNOMED-CT ontology. To address the sparseness of features in the lower levels of the SNOMED-CT hierarchy, we employed a subsumption process to generate and maintain features at higher levels of the hierarchy, ensuring the inclusion of all subordinate features.

### Feature Selection

Given the high dimensionality, with the number of symptom features exceeding 12,000, the dataset could not be analysed statistically. Instead, a genetic approach was taken. First, symptom features were removed where less than 0.5% of all patients or less than 5% of lung cancer patients had the symptom documented in their notes. Thereafter, the remaining features were ranked according to their Bayesian importance value, calculated as:

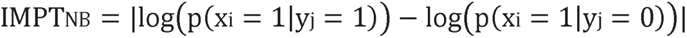

where xi and yj are 1-dimensional binary arrays indicating the presence of feature i and diagnosis j for each patient.

Following the ranking of all features, starting from the lowest ranking symptom, symptoms were removed should they have a Jaccard coefficient greater than 0.8. Thereafter, the remaining symptoms were input into a tabu asexual genetic algorithm (TAGA) [25], configured to select the feature set which maximises the area under the receiver operating characteristic curve (AUROC). TAGA was tasked with returning λ features, where λ is a number between 5 and 20. The rationale for capping the number of features included in a model at 20 was to ensure the interpretability of the final diagnostic criteria and to prevent overfitting.

### Model Development and Evaluation

20% of the data was held out of the model development process and used as a test set, with the remaining 80% being used for training and validation. For each model, a 5-fold cross-validation process was applied to select the most relevant features and identify the appropriate hyperparameters for the model, following which performance was examined using the test set. The performance of the trained models was assessed using the following diagnostic test characteristics; accuracy, sensitivity, and specificity, in addition to the calculation of the AUROC, with AUROC acting as the primary evaluation measure.

This study considered the following classification models: Logistic Regression, Mixed Naïve Bayes, and Decision Trees. The rationale for the selection of these models lies in their interpretability and ease of application.

### Comparison with existing risk tools

To determine whether the proposed method improves the diagnosis of lung cancer beyond that of existing methods that make use of similar features, a comparison with existing risk tools was performed. The proposed method was compared against the lung cancer component of the QCancer score [26,27] and the lung cancer-related risk assessment tools listed on the Cancer Research UK website [28].

In applying the QCancer score, the publicly available weights were used, and the score calculated on the same test set used for all previous comparisons. The risk assessment tools (RATs) of Hamilton et al. (2005) [28] are solely a set of feature combinations and their associated positive predictive values. Therefore, to apply the RATs to the data used in this body of work, a logistic regression model was trained for each feature combination, using the training set used for all previous experiments, returning the probability of lung cancer for each patient in the test set. Thereafter, the highest probability of all feature combinations was regarded as the final prediction for each patient. As before, the AUROC was used to compare each model.

## Results

### Demographic Information

In total, 75002 patients (35628 female) were included in this study. The study population had a mean age of 63 years ± 14 years. 36123 identified as ‘White’, 20219 identified as ‘Asian’, 7851 identified as ‘Black’, 3330 identified as ‘Other’ and 835 identified as ‘Mixed Ethnicity’. Two and 6644 patients were missing sex and ethnicity data, respectively, which were imputed.

In total, over the 12-month observation period after the first CXR, a total of 1012 lung cancer diagnoses were made. The occurrence of lung cancer at each monthly increment are shown in Fig. 1a. Also, plotted are the number of diagnoses made following a repeat scan. The total number of diagnoses following the first scan plateaued four months post-CXR, with additional diagnoses after which time being made only after a further CXR. Aside from lung cancer, other common respiratory diagnoses in the dataset included: COPD (n=1883), atelectasis (n=2432) and pneumonia (n=398).

### Risk-Score Performance Characteristics

Fig. 1b shows the performance of each of the three models, in terms of AUROC, across all 12 time intervals. The performance of the logistic regression model significantly outperformed the other two models, in terms of absolute performance but also model stability, denoted by the reduced standard deviation of AUROC. Of note, the performance of all models was less stable in the first five months, highlighting the likelihood of poorer class labelling resulting from a delay in diagnoses being uploaded to the EHRs. Considering the stabilisation in performance at five months, coupled with the plateau in diagnoses without additional scans, to strike the balance between the highest quality labelling and stable model performance, the ground truth labels established at 5-months were used for all future experiments.

### Influence of Age, Sex and Ethnicity on Risk-Score Performance

Table 1 shows the performance of the model solely using the symptoms found in the EHR of the patient, then with the inclusion of demographic data. The inclusion of age and ethnicity was shown to improve the diagnostic performance of the model, increasing AUROC to 0.69 and 0.67, respectively. Gender did not improve model performance in isolation. The inclusion of age, gender and ethnicity improved model performance across all metrics resulting in an AUROC of 0.72, with an associated sensitivity and specificity of 0.69 and 0.67, respectively.

**Table 1:** Performance characteristics of the logistic regression model on the test set, when each combination of the demographic features is incorporated. Values in brackets indicate the mean and standard deviation of the cross-validation performed on the training set.

| Input Features | Accuracy | Balanced Accuracy | AUROC | Sensitivity | Specificity |
| --- | --- | --- | --- | --- | --- |
| Symptoms Only | 0.78<br>(0.77 ± 0.02) | 0.59<br>(0.6 ± 0.01) | 0.63<br>(0.63 ± 0.02) | 0.41<br>(0.44 ± 0.03) | 0.78<br>(0.77 ± 0.02) |
| Symptoms and Age | 0.66<br>(0.66 ± 0.00) | 0.64<br>(0.66 ± 0.01) | 0.69<br>(0.71 ± 0.01) | 0.62<br>(0.66 ± 0.03) | 0.66<br>(0.66 ± 0.00) |
| Symptoms and Gender | 0.78<br>(0.73 ± 0.08) | 0.59<br>(0.6 ± 0.02) | 0.64<br>(0.64 ± 0.03) | 0.41<br>(0.46 ± 0.07) | 0.78<br>(0.74 ± 0.08) |
| Symptoms and Ethnicity | 0.54<br>(0.55 ± 0.02) | 0.61<br>(0.61 ± 0.02) | 0.67<br>(0.66 ± 0.02) | 0.69<br>(0.68 ± 0.06) | 0.54<br>(0.55 ± 0.02) |
| Symptoms, Age and Gender | 0.66<br>(0.66 ± 0.00) | 0.66<br>(0.67 ± 0.01) | 0.7<br>(0.72 ± 0.01) | 0.66<br>(0.67 ± 0.02) | 0.66<br>(0.66 ± 0.00) |
| Symptoms, Age and Ethnicity | 0.66<br>(0.66 ± 0.00) | 0.67<br>(0.66 ± 0.01) | 0.71<br>(0.72 ± 0.01) | 0.68<br>(0.67 ± 0.02) | 0.66<br>(0.66 ± 0.00) |
| Symptoms, Gender and Ethnicity | 0.66<br>(0.63 ± 0.04) | 0.6<br>(0.61 ± 0.02) | 0.68<br>(0.67 ± 0.02) | 0.54<br>(0.59 ± 0.04) | 0.66<br>(0.63 ± 0.04) |
| Symptoms, Age, Gender and Ethnicity | 0.66<br>(0.66 ± 0.00) | 0.67<br>(0.67 ± 0.02) | 0.72<br>(0.72 ± 0.01) | 0.69<br>(0.69 ± 0.03) | 0.66<br>(0.66 ± 0.00) |

### Feature Importance

To understand how each predictor influences the prediction of lung cancer, SHAP (Shapley Additive Explanations) values were calculated (Fig. 2). The most influential feature was age, with older individuals exhibiting a significant increase in the model’s output towards predicting lung cancer. Additionally, the presence of an ‘existing disorder of the lung’ was found to positively impact the prediction. Notably, individuals of white ethnicity had the greatest influence on the model outputs, increasing the SHAP value towards the prediction of lung cancer, although all ethnicities displayed varying degrees of impact toward a positive diagnosis. Males had an increased SHAP value, contributing to the prediction. Conversely, the presence of a ‘congenital anomaly of a great vessel’ and ‘disorders of the retroperitoneal compartment’ reduced the SHAP value. Context-dependent factors, such as pain, bleeding, and arthropathy, also reduced the SHAP value, making a prediction of lung cancer less likely. Finally, the presence of a cough was found to increase the SHAP value, further emphasising its relevance in the prediction of lung cancer.

**Figure 2:**
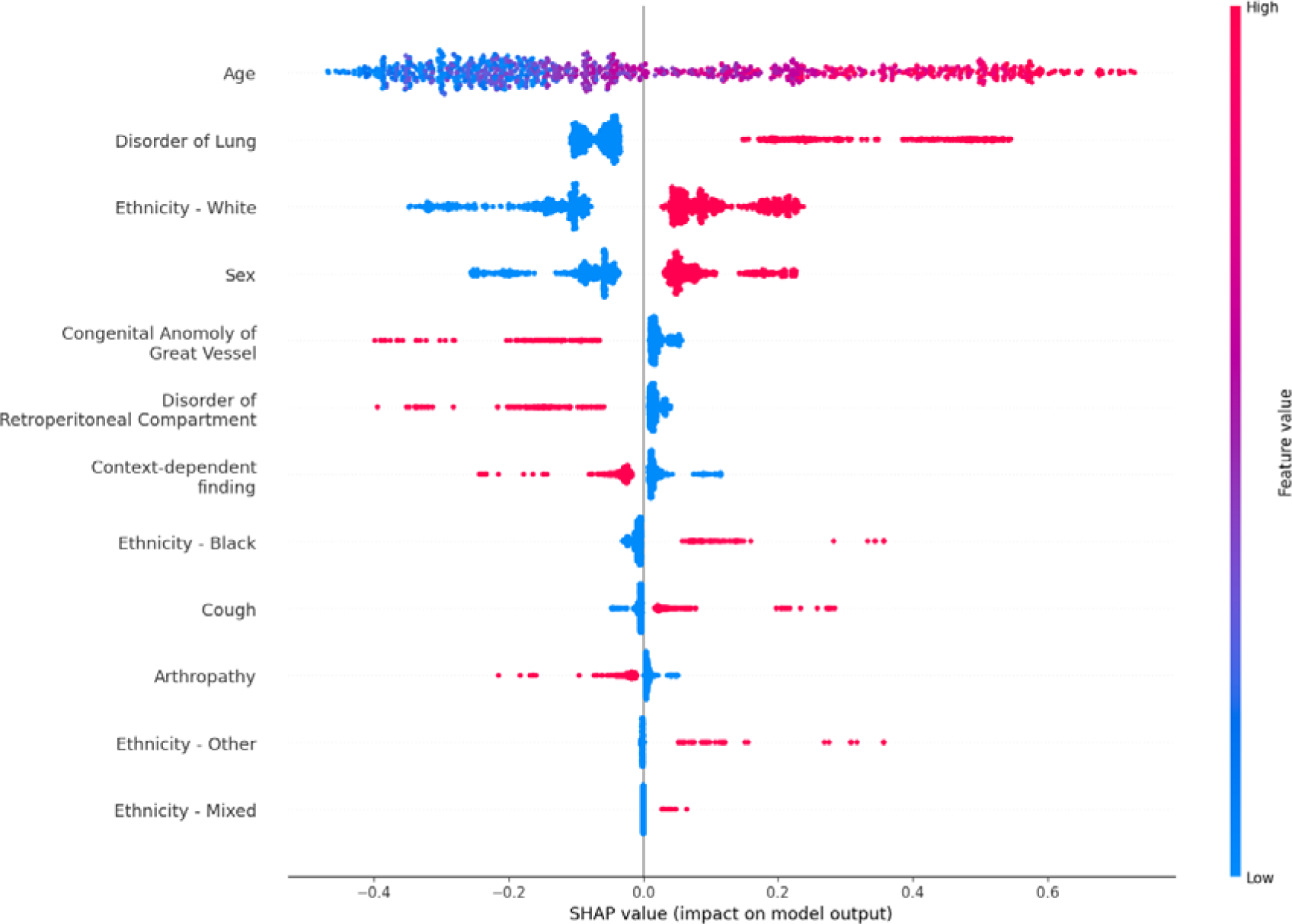
A summary plot of the SHAP values denoting the impact of each feature in the best performing model, on the prediction of lung cancer. Shading of each datapoint indicates the value of the feature. For all binary features, except age, a red value denotes a “true” value and blue denotes a false value. For age, the bluer a datapoint reflect a younger age, and the redder a data point, the older the patient.

Given the high-level nature of several the features, due to the subsumption process applied, an exploration into what symptoms or co-morbidities comprised such features was performed. Fig. 3 shows each of the selected features, and some of the most prominent features which comprise them.

**Figure 3:**
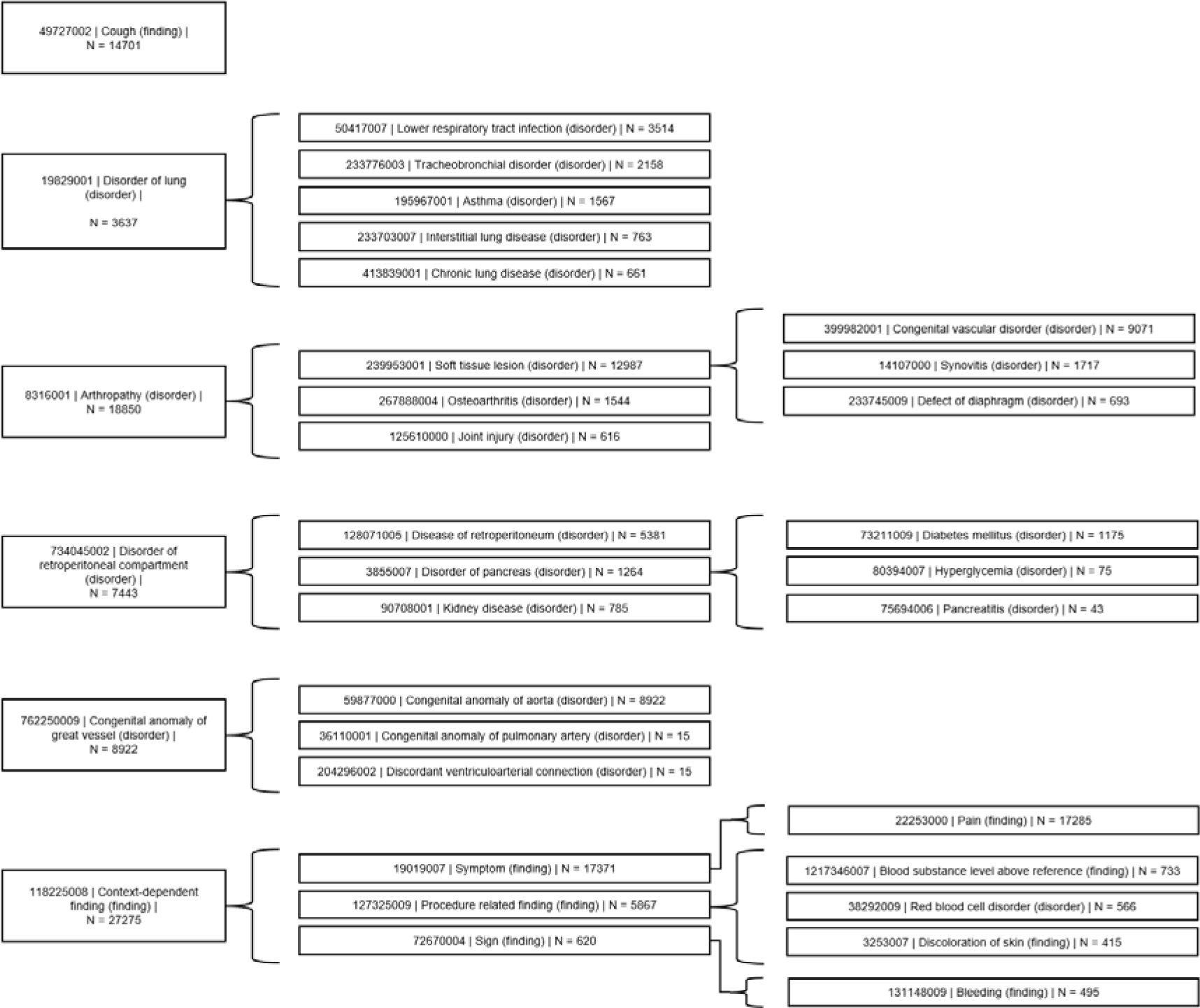
Visualisation of common concepts subsumed into higher level concepts in the SNOMED-CT hierarchy.

### Comparison of the proposed approach with other cancer risk tools

Fig. 4 shows the receiver operating characteristic curve of the model produced using the methods described in this paper, the QCancer score [26,27], and the lung cancer related risk assessment tools listed on the Cancer Research UK website [28]. As previously reported, the proposed methods resulted in an AUROC of 0.72. The application of the QCancer calculator to the test set used throughout this paper resulted in an AUROC of 0.67 and the methods of Hamilton et al. (2005) achieved and AUROC of 0.55.

**Figure 4:**
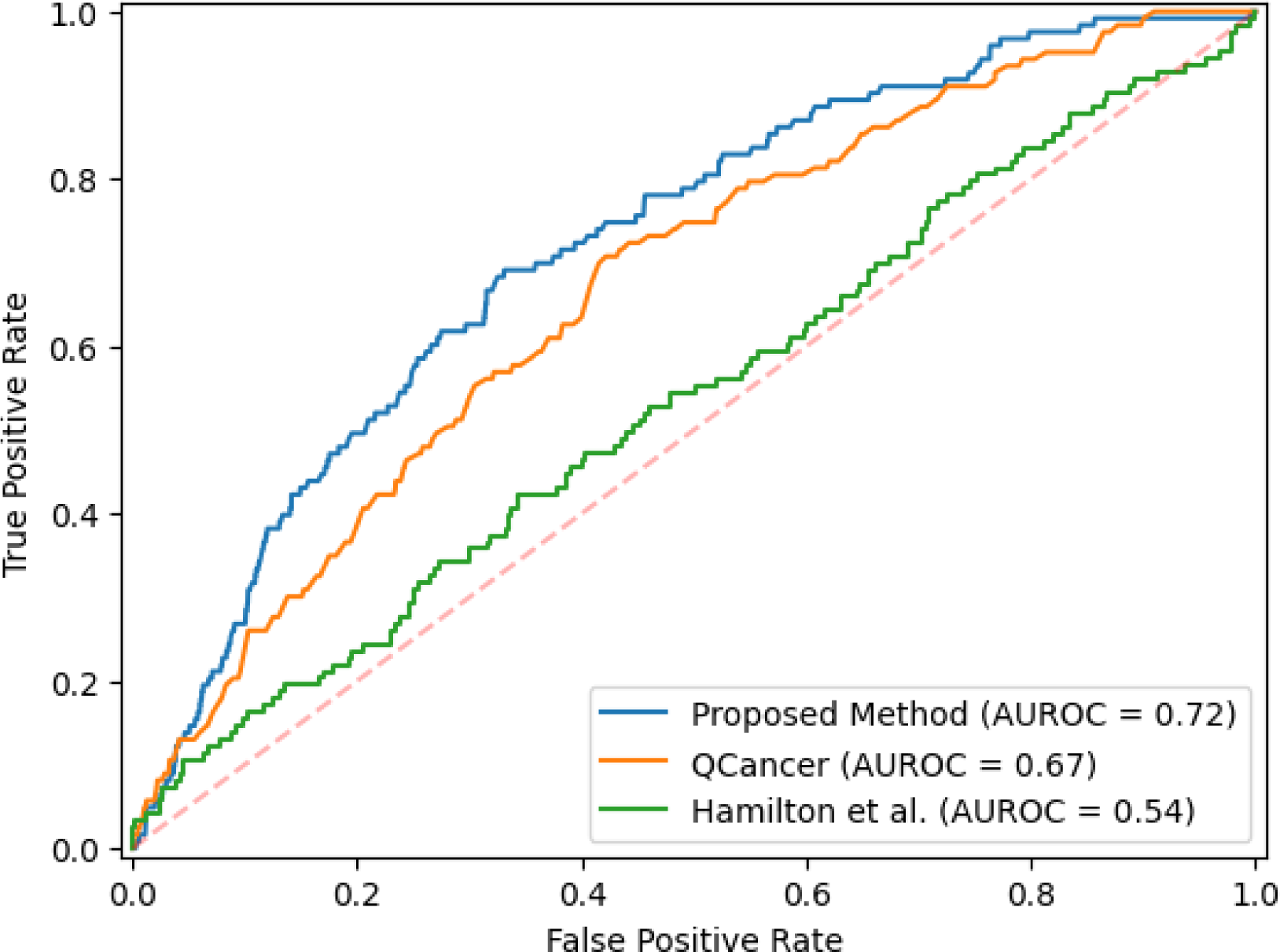
Receiver operating characteristic curves for the proposed method, QCancer and methods of Hamilton et al. (2005), when applied to the test set used in this study.

## Discussion

This work aimed to explore the use of NLP for the extraction of SNOMED-CT concepts from unstructured clinical free-text, coupled with subsumption techniques to address the challenges posed by sparse features in high-dimensional datasets. Leveraging genetic optimisation and machine learning, the generated dataset was used to develop a predictive model for lung cancer diagnosis. Model development resulted in a classifier with stable performance characterised by low standard deviations between the cross-validation folds and an AUROC of 0.72. Additionally, the model offers a balanced trade-off between sensitivity and specificity with values of 0.69 and 0.66, respectively. Notably, our proposed methodology outperforms both the QCancer calculator [26,27] and the methods introduced by Hamilton et al. (2005) [28], highlighting the promise NLP and machine learning approaches could have for the curation of rich datasets and the development of robust predictive models in the field of lung cancer risk assessment.

The incorporation of subsumption techniques helped mitigate the challenges posed by sparse features within our predictive model. By hierarchically organising and abstracting SNOMED-CT concepts, subsumption allowed us to identify broader, higher-level categories that encapsulate a range of related clinical terms. This not only alleviated the risk of overfitting and unreliable performance, a common concern in models trained on sparse data [23,24], but enhanced the generalisability of our model. However, the introduction of more abstract, top-level features meant that the final model was rooted in a level of granularity less commonly used in routine clinical practice. This has important implications for the practical translation and messaging of the model, highlighting the need for a clear and effective strategy to bridge the gap between the model’s output, which operates at a higher conceptual level, and the clinical realities on the ground, which makes use of specific and well-established terminology.

The primary function of our model is to evaluate the likelihood of a positive lung cancer diagnosis when a patient enters the clinical pathway for this purpose. While this is a valuable step in enhancing early diagnosis and intervention, the success of a diagnostic tool is often measured by its ability to identify patients even before they enter the diagnostic pathway [29], ultimately achieving a significant stage shift in the diagnostic process which is associated with improved mortality rates [30]. The primary limitation of this study is its reliance on secondary care data, which did not provide sufficient longitudinal information to facilitate such an analysis. It is essential to recognise that most patient interactions with the healthcare system before a lung cancer diagnosis occur in primary care facilities, where symptoms are first reported and initial evaluations are made [31–34]. The absence of primary care data in our study thus limits the real-world applicability of the developed methods and highlights the need for future efforts to incorporate primary care data to truly impact early detection and diagnosis in clinical practice.

A core limitation relates to documentation bias. Although purely data-driven methods were employed to derive the features predictive of lung cancer, most patients had only one document before their CXR, a referral letter. Therefore, we must consider the possibility that the referring clinician may only include symptoms that they perceive to be relevant to the suspected diagnosis for which the scan is required, omitting other symptoms which may prove predictive. Such a limitation will often be present in such predictive modelling studies. However, if each patient were to have more clinical notes before the suspecting of lung cancer the effect of such bias may be reduced.

Clinically, the absence of staging data restricts our insight into the model’s capacity to identify lung cancer at an early stage, which is crucial for understanding the impact of the predictions on patient outcomes. Additionally, the NER methods employed were not trained to extract genetic variants from pathology reports, specifically lung-cancer specific risk loci, which could further improve the performance of the model [35]. Future studies with access to more comprehensive and longitudinal patient data, including primary care information, genomic data and staging details, could help address these limitations and further enhance the efficacy and generalisability of the developed predictive model.

## Conclusions

This research highlights the potential of combining natural language processing and machine learning techniques to enhance diagnostic criteria for lung cancer using unstructured healthcare data. The study’s key findings include the successful identification of discriminating features associated with lung cancer diagnosis and achieving promising AUROC scores which outperform other comparable risk assessment tools. Such advancements hold promise for improving patient care and outcomes, albeit with a need to address certain limitations through the incorporation of primary care data for more comprehensive and unbiased criteria development.

## Data Availability

The datasets generated during and/or analysed during the current study are not publicly available due their identifiable, sensitive, and confidential nature. Data are however available from the authors upon reasonable request and with permission of Barts Health NHS Trust Information Governance Team and with appropriate approvals in place from the NHS Confidentiality Advisory Group.

## List of Abbreviations

AUROC: Area Under the Receiver Operating Characteristic curve
COPD: Chronic Obstructive Pulmonary Disease
CT: Computed Tomography
CXR: Chest X-ray
EHR: Electronic Health Record
ICD-10: International Classification of Diseases
NER: Named Entity Recognition
NHS: National Health Service
NLP: Natural Language Processing
NSCLC: Non-Small Cell Lung Cancer
SHAP: Shapley Additive Explanations
SNOMED-CT: Systematized Nomenclature of Medicine - Clinical Terms
TAGA: Tabu Asexual Genetic Algorithm

## Declarations

### Ethics Approval and Consent to Participate

This study was submitted to an NHS Research Ethics Committee, with subsequent approval being granted by the NHS Health Research Authority (IRAS ID: 320934). The requirement for informed consent to participate was waived by the NHS Health Research Authority following support, granted by the NHS Confidentiality Advisory Group, under Section 251 of the National Health Service Act 2006. The study also adhered to the NHS National Data Opt-Out to respect patients’ decision to opt out of the use of their data for research purposes.

### Consent for Publication

Not applicable

### Authors’ contributions

**AH:** Data curation, Software, Validation, Formal analysis, Project administration, Visualization, Writing – original draft; **SW:** Supervision, Project administration, Writing – original draft, Resources; **WR:** Writing – original draft**; CG:** Resources; **C T :**Funding acquisition, Project administration, Resources; **JC:** Supervision, Project administration; **All authors** : Conceptualisation, Methodology, Investigation, Writing – review & editing

## Acknowledgement

We wish to acknowledge the technical support team at Clinithink Ltd. for their assistance in developing the NLP methods employed in this study.

## References

1 Sung H, Ferlay J, Siegel RL, et al. Global Cancer Statistics 2020: GLOBOCAN Estimates of Incidence and Mortality Worldwide for 36 Cancers in 185 Countries. CA Cancer J Clin. 2021;71:209–49.

2 Case-mix Adjusted Percentage of Cancers Diagnosed at Stages 1 and 2 in England - NHS Digital. https://digital.nhs.uk/data-and-information/publications/statistical/case-mix-adjusted-percentage-of-cancers-diagnosed-at-stages-1-and-2-in-england (accessed 13 September 2023)

3 Cancer survival in England - Office for National Statistics. https://www.ons.gov.uk/peoplepopulationandcommunity/healthandsocialcare/conditionsanddiseases/bulletins/cancersurvivalinengland/stageatdiagnosisandchildhoodpatientsfollowedupto2018 (accessed 13 September 2023)

4 Reduced Lung-Cancer Mortality with Low-Dose Computed Tomographic Screening. New England Journal of Medicine. 2011;365:395–409.

5 Phadke NA, del Carmen MG, Goldstein SA, et al. Trends in Ambulatory Electronic Consultations During the COVID-19 Pandemic. J Gen Intern Med. 2020;35:3117.

6 Hill E, Mehta H, Sharma S, et al. Risk Factors Associated with Post-Acute Sequelae of SARS-CoV-2 in an EHR Cohort: A National COVID Cohort Collaborative (N3C) Analysis as part of the NIH RECOVER program. medRxiv. Published Online First: 17 August 2022. doi: 10.1101/2022.08.15.22278603

7 Prado MG, Kessler LG, Au MA, et al. Symptoms and signs of lung cancer prior to diagnosis: Comparative study using electronic health records. medRxiv. 2022;2022.06.01.22275657.

8 Wong A, Young AT, Liang AS, et al. Development and Validation of an Electronic Health Record-Based Machine Learning Model to Estimate Delirium Risk in Newly Hospitalized Patients Without Known Cognitive Impairment. JAMA Netw Open. 2018;1:e181018.

9 van Laar SA, Gombert-Handoko KB, Guchelaar HJ, et al. An Electronic Health Record Text Mining Tool to Collect Real-World Drug Treatment Outcomes: A Validation Study in Patients With Metastatic Renal Cell Carcinoma. Clin Pharmacol Ther. 2020;108:644–52.

10 Houston A, Cosma G, Turner P, et al. Predicting surgical outcomes for chronic exertional compartment syndrome using a machine learning framework with embedded trust by interrogation strategies. Scientific Reports 2021 11:1. 2021;11:1–15.

11 Naran S, Hudovsky A, Antscherl J, et al. Audit of accuracy of clinical coding in oral surgery. Br J Oral Maxillofac Surg. 2014;52:735–9.

12 Nouraei SAR, Hudovsky A, Frampton AE, et al. A Study of Clinical Coding Accuracy in Surgery: Implications for the Use of Administrative Big Data for Outcomes Management. Ann Surg. 2015;261:1096–107.

13 Koleck TA, Dreisbach C, Bourne PE, et al. Natural language processing of symptoms documented in free-text narratives of electronic health records: a systematic review. J Am Med Inform Assoc. 2019;26:364–79.

14 Weissman GE, Harhay MO, Lugo RM, et al. Natural Language Processing to Assess Documentation of Features of Critical Illness in Discharge Documents of Acute Respiratory Distress Syndrome Survivors. Ann Am Thorac Soc . 2016;13:1538–45.

15 Greenwald JL, Cronin PR, Carballo V, et al. A Novel Model for Predicting Rehospitalization Risk Incorporating Physical Function, Cognitive Status, and Psychosocial Support Using Natural Language Processing. Med Care. 2017;55:261–6.

16 Hu D, Li S, Zhang H, et al. Using Natural Language Processing and Machine Learning to Preoperatively Predict Lymph Node Metastasis for Non-Small Cell Lung Cancer With Electronic Medical Records: Development and Validation Study. JMIR Med Inform. 2022;10. doi: 10.2196/35475

17 Chase HS, Mitrani LR, Lu GG, et al. Early recognition of multiple sclerosis using natural language processing of the electronic health record. BMC Med Inform Decis Mak . 2017;17:24.

18 Zhou L, Baughman AW, Lei VJ, et al. Identifying Patients with Depression Using Free-text Clinical Documents. Stud Health Technol Inform. 2015;216:629–33.

19 Fodeh SJ, Zirkle M, Finch D, et al. MedCat: A framework for high level conceptualization of medical notes. Proceedings - IEEE 13th International Conference on Data Mining Workshops, ICDMW 2013. 2013;274–80.

20 Bean DM, Kraljevic Z, Shek A, et al. Hospital-wide natural language processing summarising the health data of 1 million patients. PLOS Digital Health. 2023;2:e0000218.

21 Lee D, de Keizer N, Lau F, et al. Literature review of SNOMED CT use. Journal of the American Medical Informatics Association. 2014;21:e11–9.

22 Benson T, Grieve G. Principles of Health Interoperability. Published Online First: 2016. doi: 10.1007/978-3-319-30370-3

23 Avanzi B, Taylor G, Wang M, et al. Machine Learning with High-Cardinality Categorical Features in Actuarial Applications. Published Online First: 30 January 2023.

24 Ohno-Machado L, Musen MA. Learning rare categories in backpropagation. Lecture Notes in Computer Science (including subseries Lecture Notes in Artificial Intelligence and Lecture Notes in Bioinformatics). 1995;991:201–9.

25 Salesi S, Cosma G, Mavrovouniotis M. TAGA: Tabu Asexual Genetic Algorithm embedded in a filter/filter feature selection approach for high-dimensional data. Inf Sci (N Y). 2021;565:105– 27.

26 Hippisley-Cox J, Coupland C. Symptoms and risk factors to identify women with suspected cancer in primary care: derivation and validation of an algorithm. Br J Gen Pract. 2013;63. doi: 10.3399/BJGP13X660733

27 Hippisley-Cox J, Coupland C. Symptoms and risk factors to identify men with suspected cancer in primary care: derivation and validation of an algorithm. Br J Gen Pract. 2013;63. doi: 10.3399/BJGP13X660724

28 Hamilton W, Peters TJ, Round A, et al. What are the clinical features of lung cancer before the diagnosis is made? A population based case-control study. Thorax. 2005;60:1059–65.

29 Balata H, Quaife SL, Craig C, et al. Early Diagnosis and Lung Cancer Screening. Clin Oncol. 2022;34:708–15.

30 Flores R, Patel P, Alpert N, et al. Association of Stage Shift and Population Mortality Among Patients With Non–Small Cell Lung Cancer. JAMA Netw Open . 2021;4:e2137508–e2137508.

31 Bradley SH, Kennedy MPT, Neal RD. Recognising Lung Cancer in Primary Care. Adv Ther. 2019;36:19–30.

32 Holtedahl K, Scheel BI, Johansen ML. General practitioners’ participation in cancer treatment in Norway. Rural Remote Health. 2018;18. doi: 10.22605/RRH4276

33 Tørring ML, Frydenberg M, Hansen RP, et al. Evidence of increasing mortality with longer diagnostic intervals for five common cancers: a cohort study in primary care. Eur J Cancer . 2013;49:2187–98.

34 Ewing M, Naredi P, Nemes S, et al. Increased consultation frequency in primary care, a risk marker for cancer: a case-control study. Scand J Prim Health Care. 2016;34:204–11.

35 Timofeeva MN, Hung RJ, Rafnar T, et al. Influence of common genetic variation on lung cancer risk: meta-analysis of 14 900 cases and 29 485 controls. Human molecular genetics, 2012;21:4980–4995.

