## Supplementary Tables for "Automated Derivation of Diagnostic Criteria for Lung Cancer using Natural Language Processing on Electronic Health Records: A pilot study"

**Supplementary File**

*Frequency Statistics for NLP Derived SNOMED-CT Concepts*

The top 100 features identified by the core problems query set and human phenotype ontology query set, ordered according to the entire patient population, are presented in tables S1 and S2, respectively.

| Table S1: Top 100 most frequently identified HPO concepts. HPO, Human Phenotype Ontology | | | |
| --- | --- | --- | --- |
| **Concept Name** | **Not Lung Cancer (N)** | **Lung Cancer (N)** | **Total Hits (N)** |
| Pain | 10151 | 98 | 27523 |
| Cough | 6191 | 69 | 17458 |
| Chest pain | 4784 | 45 | 12721 |
| Disorder of lung | 4779 | 55 | 12504 |
| Dyspnoea | 4978 | 63 | 12464 |
| Traumatic injury | 3557 | 33 | 9898 |
| Physical examination procedure | 2518 | 24 | 7133 |
| Fall | 2124 | 31 | 5581 |
| Abdominal pain | 1803 | 20 | 4893 |
| Respiratory tract infection | 1724 | 14 | 4678 |
| Hypertensive disorder systemic arterial | 1591 | 15 | 4501 |
| Pain in limb | 1574 | 9 | 4442 |
| Chronic obstructive lung disease | 1530 | 16 | 4318 |
| Lower respiratory tract infection | 1535 | 12 | 4139 |
| Fever | 1314 | 14 | 3631 |
| Tobacco user | 1110 | 17 | 3549 |
| Smoker | 1107 | 16 | 3532 |
| Fracture of bone | 1300 | 14 | 3460 |
| Musculoskeletal pain | 1153 | 4 | 3239 |
| Diabetes mellitus | 966 | 4 | 3093 |
| Disorder of skin | 1061 | 11 | 3043 |
| Pain in lower limb | 1044 | 5 | 2984 |
| Pleural effusion | 1053 | 12 | 2673 |
| Coronary artery bypass grafting | 1090 | 6 | 2619 |
| Skin lesion | 858 | 7 | 2514 |
| Asthma | 764 | 10 | 2381 |
| Asthma without status asthmaticus | 763 | 10 | 2378 |
| Backache | 792 | 6 | 2295 |
| Osteoarthritis | 762 | 3 | 2159 |
| Vomiting | 797 | 7 | 2118 |
| Oedema | 778 | 7 | 2052 |
| Viral disease | 690 | 6 | 2003 |
| Atrial arrhythmia | 726 | 8 | 1995 |
| Atrial fibrillation | 668 | 7 | 1858 |
| Joint pain | 645 | 1 | 1796 |
| Diabetes mellitus type 2 | 557 | 2 | 1758 |
| Heart failure | 637 | 3 | 1693 |
| Disorder of brain | 553 | 5 | 1607 |
| Pain in upper limb | 582 | 4 | 1597 |
| Cerebrovascular disease | 579 | 3 | 1582 |
| Wheezing | 532 | 4 | 1578 |
| Kidney disease | 514 | 5 | 1528 |
| Pneumonitis | 599 | 7 | 1522 |
| Inflammatory dermatosis | 535 | 5 | 1498 |
| Sepsis | 564 | 6 | 1472 |
| Weight decreased | 495 | 5 | 1460 |
| Disorder of artery | 531 | 5 | 1459 |
| Arthritis | 507 | 5 | 1437 |
| Clouded consciousness | 551 | 4 | 1375 |
| Dizziness | 490 | 8 | 1323 |
| Caesarean section | 507 | 2 | 1319 |
| Pneumonia | 486 | 7 | 1291 |
| Cerebrovascular accident | 469 | 1 | 1269 |
| Myocardial infarction | 460 | 4 | 1221 |
| Muscle disorder | 512 | 4 | 1195 |
| Hypercholesterolemia | 415 | 4 | 1190 |
| Injury of head | 384 | 4 | 1183 |
| Duodenal ulcer disease | 453 | 6 | 1116 |
| Headache | 361 | 4 | 1107 |
| Renal impairment | 367 | 4 | 1099 |
| Fracture of rib | 434 | 4 | 1096 |
| Mass of body structure | 423 | 6 | 1090 |
| Allergic disposition | 380 | 2 | 1052 |
| Haemoptysis | 353 | 1 | 1043 |
| Laceration injury | 362 | 1 | 1021 |
| Neuropathy | 356 | 3 | 988 |
| Chest wall pain | 321 | 0 | 973 |
| Pneumothorax | 395 | 4 | 953 |
| Palpitations | 381 | 3 | 932 |
| Dyspnoea on exertion | 342 | 2 | 929 |
| Epigastric pain | 333 | 3 | 878 |
| Hyperlipidaemia | 316 | 1 | 875 |
| Peripheral vascular disease | 333 | 3 | 867 |
| Ex smoker | 266 | 0 | 861 |
| Disorder of the peripheral nervous system | 302 | 4 | 859 |
| Pain in pelvis | 277 | 1 | 851 |
| Urinary tract infectious disease | 295 | 4 | 847 |
| Fatigue | 305 | 4 | 812 |
| Peripheral nerve disease | 291 | 1 | 800 |
| Allergy to drug | 281 | 2 | 799 |
| Nausea | 299 | 4 | 788 |
| Anaemia | 280 | 0 | 787 |
| Chronic kidney disease | 237 | 3 | 760 |
| Scoliosis deformity of spine | 337 | 3 | 759 |
| Hiatal hernia | 322 | 2 | 739 |
| Acute myocardial infarction | 305 | 2 | 738 |
| Tachycardia | 302 | 0 | 731 |
| Mood disorder | 265 | 2 | 718 |
| Disorder of thyroid gland | 235 | 3 | 711 |
| Altered bowel function | 244 | 2 | 705 |
| Shoulder pain | 251 | 3 | 689 |
| Sleep disorder | 228 | 3 | 681 |
| Thrombosis | 271 | 2 | 679 |
| Bronchiectasis | 253 | 2 | 678 |
| Low back pain | 218 | 2 | 667 |
| Knee pain | 221 | 1 | 652 |
| Seizure | 208 | 1 | 649 |
| Depressive disorder | 235 | 1 | 646 |
| Diarrhoea | 218 | 2 | 600 |
| Constipation | 217 | 1 | 597 |

| Table S2: Top 100 most frequently identified HPO concepts. HPO, Human Phenotype Ontology | | | |
| --- | --- | --- | --- |
| **Concept Name** | **Not Lung Cancer (N)** | **Lung Cancer (N)** | **Total Hits (N)** |
| Abnormality of the thorax | 11479 | 107 | 30088 |
| Pain | 10145 | 108 | 27555 |
| Abnormality of the respiratory system | 10103 | 85 | 26690 |
| Abnormality of the cardiovascular system | 6865 | 69 | 18420 |
| Cough | 6194 | 56 | 17444 |
| Breathing dysregulation | 5649 | 51 | 14322 |
| Respiratory distress | 5644 | 50 | 14311 |
| Abnormality of lung morphology | 5320 | 45 | 13923 |
| Chest pain | 4785 | 50 | 12748 |
| Dyspnoea | 4993 | 46 | 12452 |
| Abnormality of the mediastinum | 4576 | 44 | 12130 |
| Abnormal heart morphology | 4490 | 45 | 11891 |
| Abnormality of head or neck | 3098 | 25 | 8673 |
| Abnormality of connective tissue | 2907 | 34 | 7554 |
| Abnormality of the head | 2659 | 24 | 7343 |
| Abnormality of the digestive system | 2648 | 24 | 6941 |
| Abnormality of the gastrointestinal tract | 2648 | 24 | 6941 |
| Abnormality of limbs | 2337 | 31 | 6619 |
| Pneumothorax | 2282 | 19 | 5616 |
| Abnormality of the tracheobronchial system | 1874 | 18 | 5310 |
| Abnormality of the bronchi | 1833 | 16 | 5193 |
| Abdominal pain | 1830 | 22 | 4960 |
| Abnormal joint morphology | 1717 | 20 | 4839 |
| Atelectasis | 1918 | 14 | 4751 |
| Arthropathy | 1668 | 20 | 4700 |
| Respiratory tract infection | 1725 | 7 | 4677 |
| Abnormality of the skeletal system | 1770 | 22 | 4671 |
| Hypertension | 1604 | 16 | 4538 |
| Abnormality of the vasculature | 1602 | 19 | 4458 |
| Limb pain | 1572 | 18 | 4439 |
| Cardiomegaly | 1757 | 16 | 4380 |
| Chronic obstructive pulmonary disease | 1534 | 14 | 4315 |
| Abnormality of the nervous system | 1503 | 13 | 4314 |
| Abnormality of the lower limb | 1493 | 18 | 4225 |
| Recurrent lower respiratory tract infections | 1541 | 7 | 4143 |
| Abnormality of the endocrine system | 1266 | 15 | 3912 |
| Fever | 1319 | 8 | 3635 |
| Abnormality of the orbital region | 1276 | 18 | 3387 |
| Morphological abnormality of the central nervous system | 1166 | 6 | 3355 |
| Abnormality of carbohydrate metabolism or homeostasis | 1010 | 14 | 3253 |
| Abnormality of the integument | 1169 | 11 | 3224 |
| Abnormality of the eye | 1205 | 16 | 3188 |
| Arthralgia or arthritis | 1139 | 16 | 3165 |
| Abnormality of the skin | 1103 | 11 | 3082 |
| Diabetes mellitus | 950 | 13 | 3081 |
| Abnormality of the genitourinary system | 1091 | 17 | 3069 |
| Lower limb pain | 1044 | 11 | 2989 |
| Arrhythmia | 1040 | 7 | 2900 |
| Rales | 960 | 11 | 2761 |
| Abnormality of the intestine | 1040 | 9 | 2699 |
| Pleural effusion | 1056 | 7 | 2672 |
| Abnormality of the urinary system | 929 | 15 | 2660 |
| Reduced consciousness or confusion | 984 | 6 | 2496 |
| Abnormality of long bone morphology | 954 | 13 | 2493 |
| Fractures of the long bones | 903 | 12 | 2427 |
| Asthma | 762 | 8 | 2372 |
| Back pain | 793 | 8 | 2317 |
| Behavioural abnormality | 881 | 6 | 2295 |
| Abnormality of the anterior segment of the globe | 923 | 11 | 2292 |
| Abnormality of cardiac atrium | 835 | 4 | 2278 |
| Abnormality of the upper limb | 774 | 11 | 2187 |
| Vomiting | 788 | 9 | 2118 |
| Supraventricular arrhythmia | 775 | 3 | 2113 |
| Arthralgia | 736 | 9 | 2040 |
| Primary atrial arrhythmia | 735 | 2 | 1994 |
| Oedema | 755 | 2 | 1989 |
| Osteoarthritis | 652 | 5 | 1876 |
| Atrial fibrillation | 676 | 2 | 1857 |
| Productive cough | 646 | 2 | 1760 |
| Abnormality of the uvea | 718 | 11 | 1748 |
| Abnormality of the pleura | 716 | 9 | 1747 |
| Type II diabetes mellitus | 535 | 10 | 1741 |
| Autoimmunity | 610 | 4 | 1735 |
| Congestive heart failure | 632 | 8 | 1688 |
| Abnormal axial skeleton morphology | 679 | 7 | 1658 |
| Abnormality of brain morphology | 556 | 1 | 1632 |
| Encephalopathy | 556 | 1 | 1631 |
| Wheezing | 550 | 3 | 1629 |
| Abnormal myocardium morphology | 603 | 8 | 1597 |
| Upper limb pain | 578 | 8 | 1587 |
| Inflammatory abnormality of the skin | 580 | 3 | 1565 |
| Nephropathy | 516 | 8 | 1541 |
| Abnormality of the kidney | 515 | 8 | 1539 |
| Abnormality of limb bone morphology | 547 | 6 | 1502 |
| Abnormality of limb bone | 540 | 6 | 1483 |
| Sepsis | 558 | 9 | 1467 |
| Abnormality of the stomach | 603 | 5 | 1466 |
| Weight loss | 499 | 2 | 1462 |
| Confusion | 558 | 4 | 1388 |
| Abnormality of the face | 515 | 5 | 1382 |
| Arthritis | 485 | 7 | 1380 |
| Inflammatory abnormality of the eye | 536 | 6 | 1341 |
| Vertigo | 493 | 5 | 1324 |
| Abnormality of the ribs | 522 | 4 | 1324 |
| Caesarean section | 504 | 6 | 1320 |
| Secondary Caesarean section | 504 | 6 | 1320 |
| Abnormality of the rib cage | 523 | 5 | 1308 |
| Abnormality of the vertebral column | 487 | 7 | 1286 |
| Stroke | 456 | 5 | 1284 |
| Pneumonia | 485 | 5 | 1273 |
